# Fine Needle Aspiration Versus the CytoCore® Motorized Rotating Needle Device for Thyroid Nodule Biopsies: A Retrospective Cohort Study

**DOI:** 10.1101/2023.09.09.23294856

**Authors:** Adarsh Verma, Rhonda McDowell, Anthony Porreca

**Author notes:** **Corresponding Author**: Adarsh Verma, MD, Radiology Associates of Clearwater, 1106 Druid Road S., Suite 302, Clearwater, FL 33756. **Funding** Funding for this study, including data collection, statistical analysis and medical writing, was provided by Praxis Medical, LLC, Tampa, Florida.

## Abstract

**Background:** While ultrasound guided fine needle aspiration (US-FNA) is commonly used to biopsy suspicious thyroid nodules, its use is associated with a high rate of nondiagnostic and indeterminate samples. The objective of this study was to compare the diagnostic performance of a new motorized FNA device (CytoCore®, Praxis Medical) to a historical cohort of patients biopsied using US-FNA within the same health system and literature controls.

**Materials and Methods:** Data from 120 patients with suspicious thyroid nodules undergoing thyroid biopsy with a motorized FNA device between March 2022 and August 2023 was retrospectively analyzed. Patient demographics, lesion characteristics, number of passes required, Bethesda category, and cellularity scores were compared to a historical control cohort of 100 patients who underwent US-FNA between March 2019 and March 2020. All patients underwent the procedure within the same health system. Nondiagnostic and indeterminate samples rates for the motorized FNA device were separately compared to literature controls.

**Results:** A significantly reduced median number of passes were required with the motorized FNA device compared to US-FNA (*z* = 8.235, *p* < .001). Adequate samples were obtained after the first pass for 58% of nodules biopsied with motorized FNA device compared to only 11% with US-FNA. The cumulative percentage of adequate samples increased to 98% after two passes for the motorized FNA group versus only 58% for the US-FNA group. Eleven percent of subjects in the US-FNA required 5 passes to obtain an adequate sample. The mean cellularity score was also greater for the motorized FNA group (3.5 ± 0.5 vs. 2.0 ± 0.6; *z* = 5.201, *p* < .001). Compared to published rates, the motorized had a lower nondiagnostic rate (2.0% vs. 10% to 15%) and lower indeterminate rate (8.3% vs. 20%; p=0.05) compared to the use of FNA.

**Conclusion:** The motorized FNA device requires less passes to obtain an adequate biopsy than US-FNA, thus decreasing procedure length, tissue trauma, and damage to the specimen damage. Its use is also associated with obtaining samples with a higher cellularity and lower nondiagnostic and indeterminate sample rates.

## INTRODUCTION

Thyroid cancer is the most common malignancy of the endocrine system with over 43,0000 new cases of thyroid cancer diagnosed in the U.S. in 2020.[1] Developments in imaging modalities have contributed to the increased discovery of thyroid nodules that were previously undetectable and it is now estimated that between 60 to 70% of all adults have at least one thyroid nodule. [2,3] Despite this high prevalence, most thyroid nodules are ultimately benign with malignancy rates as low as 5%. [2] Fine needle aspiration (FNA) is considered the gold standard in differentiating benign and malignant nodules due to its high specificity and sensitivity.[4-6] FNA is a minimally invasive procedure that poses low risk to the patient compared with other biopsy techniques such as core biopsy. Despite these advantages, FNA is limited by rate a nondiagnostic biopsy rate of 10% to 15% and an indeterminate result rate of 20%. [7] This leads to the need for repeat procedures in 50% to 63% of patients for whom nondiagnostic samples were obtained [8] and partial or total thyroidectomy for patients with Bethesda 3 and 4 nodules that are deemed suspicious by molecular testing. Of thyroid nodules deemed suspicious by molecular testing, 33.1% were benign, 24.4% follicular or Hurthle cell adenoma and 42.2% were malignant on the final pathology report following surgical resection. [9] Rates of thyroid surgery-specific postoperative complications, including recurrent laryngeal nerve injury, permanent hypoparathyroidism, and postoperative hematoma, range from 0.4 to 7.4% at high volume centers with population-based study reporting that this rate can be as high as 12.3%. [7]. Nondiagnostic and indeterminate findings also result in increased patient anxiety as their biopsy will need to be either repeated or the patient may need to undergo thyroid surgery. [10]

Several factors can impact sample adequacy during standard FNA including the skill and knowledge of the individual performing the biopsy, number of passes, preparation of the smears, amount of suction, capillary technique, needle diameter and the nature of the nodule itself (size, composition, vascularity). [10-12] While the introduction of rapid on-site evaluation (ROSE) has been reported to reduce the nondiagnostic rate in several studies by providing immediate feedback on sample quality, [13] a recent report indicates that only 26% of FNA samples are analyzed using ROSE due in part to a limited number of cytology technicians available to perform this analysis. [14] ROSE has also not shown the ability to significantly reduce indeterminate diagnoses or the need for repeat procedures and surgeries. [15] Using current techniques, a minimum of 3 to 4 passes are necessary to achieve adequate sampling of thyroid nodules for diagnostic purposes, with higher numbers likely required in a non-academic or community setting. [11, 12, 15, 16]

The motorized rotating fine needle thyroid biopsy device needle (CytoCore®, Praxis Medical, Tampa, Florida, USA) was developed to reduce the rate of nondiagnostic and indeterminate samples to increase the cellular yield per pass, thereby reducing the number of passes required to obtain adequate samples (Figure 1). The device is designed to reduce sample variability by enabling a more consistent sampling technique as a result of a rotational drilling action being combined with the standard in and out motion used to access the nodule with a needle. The rotation of the needle enables its beveled tip to collect a higher quantity of intact cellular material, which is optimal for determining adequacy and, ultimately, for making a diagnosis. The device, which received 501k clearance from the FDA on March 31, 2020, is ergonomically designed and enables users to operate it using one hand with no special training required.

**Figure 1.**
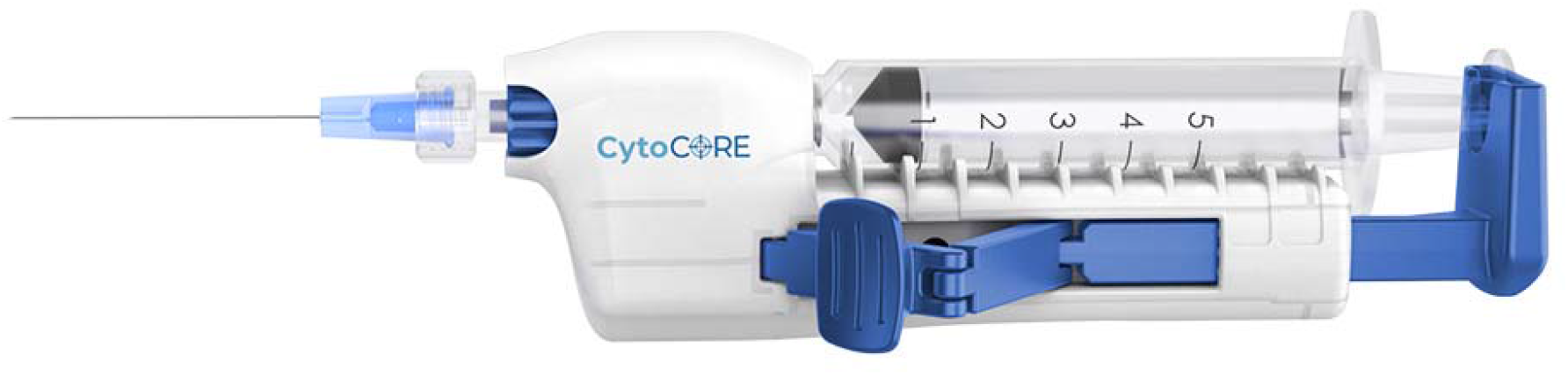
The CytoCore® motorized, rotating fine needle biopsy device.

The objective of the present study was to compare the use of the motorized FNA device to standard FNA performed using ultrasound guidance to compare number of passes required to obtain an adequate sample and the nondiagnostic and indeterminate sample rates associated with it use.

## MATERIALS & METHODS

The protocol for this retrospective study received approval under Expedited Review with waiver of consent by the BayCare Health System Institutional Review Board (FWA 00006065 IORG 0003355). A chart review was conducted to obtain data for a consecutive group of patients who underwent ultrasound-guided fine needle aspiration (US-FNA) thyroid biopsies between March 2019 and March 2020 (historical cohort) and those who had the same procedure with a motorized FNA device between March 2022 and August 2023. Patient and lesion characteristics, procedure information and pathology results were obtained and analyzed. Comparative cellularity scoring was performed by an independent cytologist comparing 20 randomly selected biopsies from both the US-FNA and the motorized FNA groups which used the same ROSE team. Since the nondiagnostic and indeterminate sample rates were not available for the historical cohort, comparisons were made based on the rates reported in the literature and recent guidelines

Nine different physicians with between 4 and 39 years of experience performed biopsies for patients in the study. All US-FNA and motorized FNA biopsies were performed using a 22-gauge x 1½ inch needle (Becton Dickinson, Franklin Lakes, New Jersy, USA) attached to the motorized biopsy device. As part of the standard of care at this institution, a cytotechnologist was present for all procedures and performed ROSE on the smears to determine if the sample met the criteria for adequacy.

### Statistical Analyses

Descriptive statistics are reported as means with standard deviations for continuous variables and as frequencies and percentages for categorical variables. Difference in proportions tests were used to compare percentage adequate sampling associated with the number of passes, cellularity scores, and the percentage of nondiagnostic, indeterminant, and determinant diagnosis classifications by the Bethesda system to results reported in the literature utilizing conventional US-FNA techniques. A p-value<0.05 was considered statistically significant.

## RESULTS

A total of 220 thyroid biopsies were included in the study with 120 patients including in the motorized FNA group and 100 patients included in the US-FNA control cohort (Table 1). The mean patient age was 62.1 ± 13.4 years for the motorized FNA group and 66.7 ± 11.1 years for the control cohort. Despite the similarity in age, the control group was significantly older (*t*(214.785) = 2.767, *p* < .01). Thyroid nodule size was larger for the FNA group compared to the motorized FNA group (2.5 ± 0.9 vs. 2.2 ± 0.9; *t*(218) = 2.203, *p* < .05). There was no significant difference for the percentage of females, nodule location or nodule appearance on ultrasound.

**Table 1.** Patient and nodule characteristics.

|  | Motorized FNA group | Control FNA group |
| --- | --- | --- |
| Average age; years ± SD | 65 | 67 |
| Female | 67% | 71% |
| Number of nodules | 120 | 100 |
| Nodule size; cm ± SD (range) | 2.2 ± 0.9<br>(1 – 7.8) | 2.5 ± 0.9<br>(1.1 – 4.4) |
| Thyroid nodule location, number (%) |  |  |
| Right lobe | 56 (46.7%) | 43 (43.0%) |
| Left lobe | 47 (39.2%) | 42 (42.0%) |
| Isthmus | 17 (14.2%) | 15 (15.0%) |
| Appearance on ultrasound |  |  |
| Hypoechoic | 21 (17.5%) | 12 (12.0%) |
| Isoechoic | 10 (8.3%) | 2 (2.0%) |
| Hyperechoic | 6 (5%) | 7 (7.0%) |
| Complex | 34 (28.3%) | 16 (16.0%) |
| Solid | 48 (40%) | 50 (50.0%) |

FNA = fine needle aspiration; SD = standard deviation

The biopsies were performed by 9 different operators, with between 4 and 39 years of clinical experience. All 120 motorized biopsies were performed using a 22 gauge /1 ½ inch BD (Benton Dickinson) needle attached to the motorized biopsy device. As part of the standard of care within this health system, a cytotechnologist was present for all procedures and performed ROSE on the smears to determine if the sample met the criteria for adequacy.

### Cytological Adequacy

Significantly fewer passes were required with the motorized FNA device with 58% of samples being adequate following first pass versus 11% for the US-FNA, and 98% adequacy following a second pass compared to 58% of patients for the control cohort (Figure 2). Eleven percent of nodules in the US-FNA group required 5 passes to obtain an adequate sample. A Mann-Whitney *U* test indicated a significantly reduced median number of passes required with the motorized FNA device, *z* = 8.235, *p* < .001.

**Figure 2.**
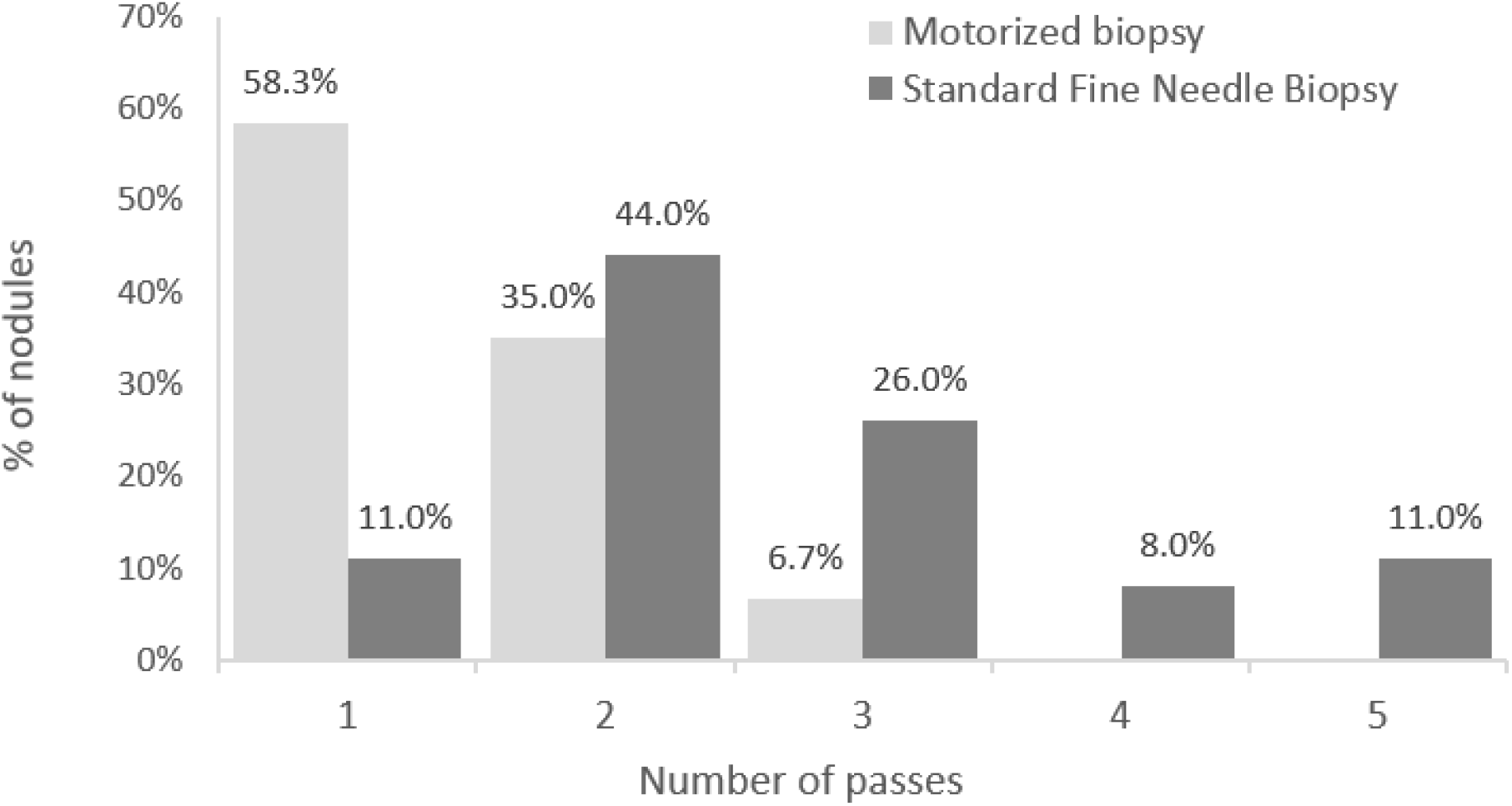
Number of passes required to obtain adequate biopsy

**Figure 3.**
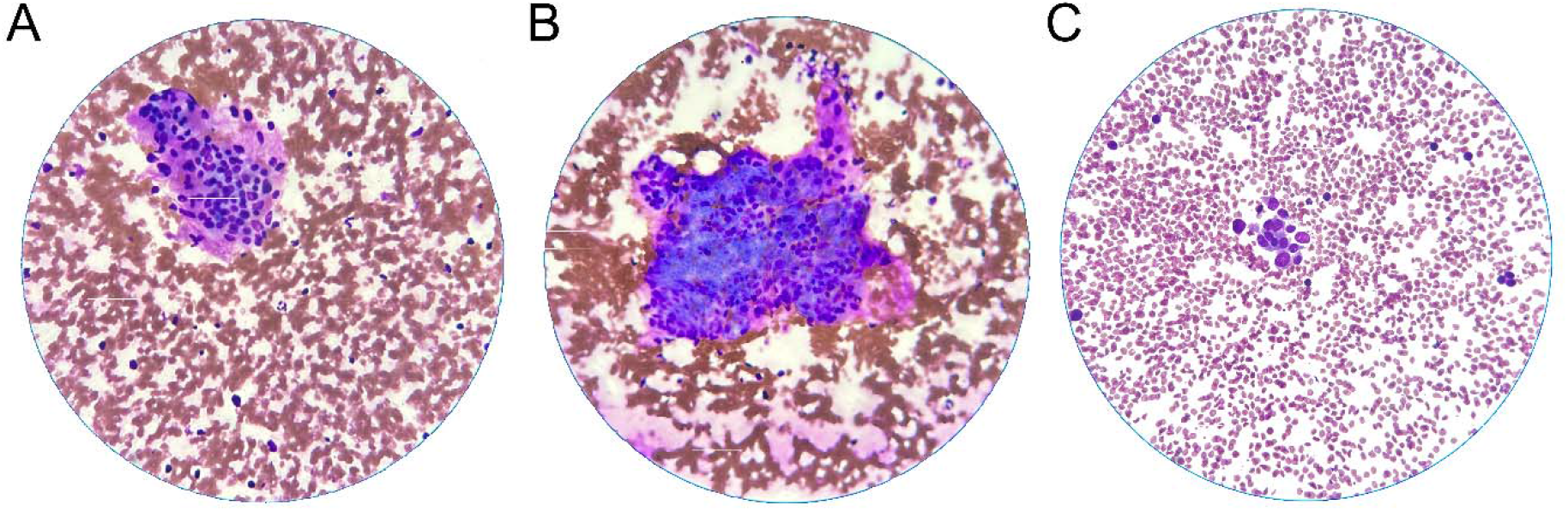
Representative cytology samples. (A) and (B) samples obtained using the motorized FNA device showing high cellularity. (C) sample obtained using standard FNA showing low cellularity.

The mean cellularity score was also greater for the motorized FNA group (3.5 ± 0.5 vs. 2.0 ± 0.6; *z* = 5.201, *p* < .001). Compared to published rates, the motorized had a lower nondiagnostic rate (2.0% vs. 10% to 15%) and lower indeterminate rate (8.3% vs. 20%; p = 0.05) compared to the use of FNA. (Table 2).

**Table 2.** Comparison of Cellularity Scores.

| Cellularity Scoring | Motorized FNA group<br>(n=20) | Control FNA group<br>(n=20) |
| --- | --- | --- |
| 0: Non-diagnostic | 0 | 0 |
| 1: Scant cellularity | 0 | 4 |
| 2: Low cellularity | 0 | 13 |
| 3: Moderate cellularity | 11 | 3 |
| 4: High cellularity | 9 | 0 |

### Pathology Diagnosis

Table 3 reports the Bethesda System classification for the 120 nodules included in the motorized FNA group. Cumulatively, pathology was able to make determinant diagnoses for 110 of the 120 (91.6%) of the samples. This included 103 (85.8%) samples identified as being benign and 7 (5.8%) noted as being malignant. Only 2 (1.7%) samples were reported as being nondiagnostic. This non-diagnostic rate is significantly lower than the published 10% to 15% nondiagnostic rate for thyroid biopsies using the standard FNA technique as noted in recently published guidelines from American Association of Endocrine Surgeons [9]. The 8.3% rate of indeterminant classifications (Bethesda Category III and IV) is also lower than the 20% that been consistently reported the same guidelines. [9]

**Table 3.** Bethesda category for samples obtained with motorized FNA device.

| Bethesda category | Number of samples (%) |
| --- | --- |
| 1: Nondiagnostic | 2 (1.7) |
| 2: Benign | 103 (85.5) |
| 3: atypia of undetermined significance or follicular lesion of undetermined significance | 7 (5.8) |
| 4: follicular neoplasm or suspicious for follicular neoplasm | 3 (2.5) |
| 5: suspicious for malignancy | 4 (3.3) |
| 6: malignant | 1 (0.8) |

## DISCUSSION

Thyroid cancer rates continue to rise across the world which has led to an increase in the number of thyroid biopsies being performed annually. The present study demonstrated that the use of a motorized device to biopsy thyroid nodules resulted in a need for fewer passes to obtain adequate cellular specimens for diagnosis when compared the use of a standard US-FNA approach. The reduced number of passes lowers tissue trauma and the risk of bleeding complications by decreasing the number of needle punctures to achieve an adequate sample and substantially improves the diagnosis of thyroid nodules as benign or malignant. The reduced trauma also likely decreases damage to the tissue sample itself, increasing the quality of the sample.

The nondiagnostic and indeterminate sample rates achieved with this device were also significantly lower than the rates reported in the American Association of Endocrine Surgeons guidelines for the definitive surgical management of thyroid disease in adults.[7] The management of an initially indeterminant biopsy can range from performing repeat US-FNA for samples categorized as Bethesda Category I, III and IV to lobectomy or thyroidectomy for those classified as Bethesda Categories IV and V. [7, 18] The actual risk of malignancy can be as low as 1 to 15% for Categories I and III, but as high as 75% for Category V. [18] Therefore, initially indeterminant diagnostic findings can result in unnecessary procedures and increased costs to the healthcare system. From a patient perspective, false positives can result in unnecessary procedures and their associated risks, and false negatives or indeterminant results can lead to delays in diagnosis and treatment.

The 2023 national average Medicare hospital and physician payment rates for performing a thyroid biopsy using US-FNA (CPT 60100) are $649 and $77, respectively, plus the cost of cytologic examination. The 2023 payment rates are $5,212 and $719, respectively, for performing a partial thyroid lobectomy (CPT 60210) and $3,600 for thyroid molecular testing (PLA code 0026U). While the motorized FNA device increases the cost of performing this procedure compared to the standard approach, it offers a significant savings by reducing the need for repeat FNA for nondiagnostic samples and possibly the need for molecular testing by lowering non-diagnostic rates due to robust sample cellularity. There is potential for even greater cost saving associated with the potential avoidance of surgical procedure. With approximately, 600,000 thyroid US-FNAs performed in the U.S. each year and reduction in nondiagnostic samples rates using the motorized FNA device from 10% to 15% to the 1.7% rate reported in the present study, we estimate that the use of this device would save the healthcare system an estimated $62 million per year as a result of a reduction in repeat FNA procedures and $210 million per year resulting from the reduced cost for molecular testing. Additionally, the health system would experience savings from a reduction in unnecessary surgical procedures. The avoidance of surgical intervention would also have a significant impact of patient’s quality of life due to potential surgical complications, vocal cord palsy, lifetime hormonal replacement, and cosmetic scarring associated with these procedures. [7]

There are several limitations associated with the present study. The retrospective nature of the study design limits the ability to make definitive conclusions from the results presented. The inclusion of data from a single health system with no prospectively defined inclusion criteria for patients creates the potential for treatment bias. The unavailability of nondiagnostic and indeterminate sample rates for the historical US-FNA control group and reliance of published rates from the literature is also an additional limitation. Prospective, randomized clinical studies across a variety of use settings with and without ROSE will be important to fully understand the potential advantages of using the motorized biopsy device. This should include the collection of longitudinal data reporting the use of molecular testing and need for surgical procedures with associated complications. While multiple physicians performed biopsies for the patients enrolled in the study and could have impacted the results, the high cellularity scoring of the samples obtained with the motorized FDA device versus standard US-FNA suggests a significant technological advantage with more diagnostic specimens during procedure. These results also suggest device performance is not related to operator experience.

Although the focus of the present study was on performance of the motorized device for the biopsy of thyroid nodules, it should be noted that the device can also be used in any soft tissue biopsy. Future studies examining these other applications in soft tissues such as lymph nodes, lung, or liver are forthcoming.

## CONCLUSION

The results of this study showed that using the motorized biopsy device improves clinical efficiency by reducing the number of passes required to obtain an adequate sample, reducing the indeterminate and nondiagnostic rates, and yielding more cellular samples compared to standard FNA. The lower number of passes reduces tissue trauma and the risk of biopsy related bleeding and infectious complication. The increase in the adequacy of samples substantially improves the ability to diagnosis the thyroid nodules as being benign or malignant. As a result of the lower nondiagnostic and indeterminate rates with motorized biopsy device, there is a reduced need for further interventions including repeat biopsies, molecular testing, and surgical procedures, resulting in potential cost savings to the health system and improved patient quality of life.

## Data Availability

All data produced in the present study are available upon reasonable request to the authors.

## Acknowledgements

We thank the following institutions and individuals who also contributed patients to this study. Morton Plant Hospital, Clearwater, FL, Mease Countryside Hospital, Countryside, FL, Mease Dunedin Hospital, Dunedin, FL, and Alexa Alsina, PA-C, Derek Samuels MD, and Brad White MD from Radiology Associates of Clearwater, Clearwater, FL. Statistical analyses were performed by DK Statistical Consulting Inc., Houston, TX. Medical writing assistance was provided by Larry Yost (The Atticus Group, LLC, Portsmouth, NH).

